# Assessment of the Prevalence of Pulmonary Tuberculosis Patients at Nakfa Hospital from 2014-2019, Eritrea

**DOI:** 10.1101/2020.02.11.20021279

**Authors:** Yafet Kesete

**Author notes:** Correspondence should be addressed to Yafet Kesete.

## Abstract

**Background:** Tuberculosis, an infectious disease, is one of the top 10 causes of death, and the leading cause from a single infectious agent M. tuberculosis. According to recent WHO estimate report, at least 3100 new TB cases occur every year in Eritrea. However, very little information is available in Eritrea related to area specific prevalence of tuberculosis and risk factors attributed to it. This is a retrospective study aimed to assess the prevalence of tuberculosis disease among patients attending at Nakfa Hospital, Eritrea.

**Methods:** A retrospective study was conducted on patients visiting Nakfa Hospital, 221km north of Asmara, Eritrea from April 2014 to March 2019. Data was extracted from secondary data sources like patient admission records and laboratory registers at Nakfa Hospital.

**Results:** A total of 1100 patients were examined for tuberculosis using acid fast staining test. The overall prevalence of smear positive pulmonary TB cases was 7.8% (86 cases out of 1100). Females (8.2%) were more prone to have a positive Tuberculosis smear than males (7.4%). According to severity of infection, 38(3.5%) of subjects were +1 positive, whereas 23(2.1%) and 24(2.2%) of patients were +2 and +3 positives respectively. The highest prevalence of pulmonary TB was observed in the adult age group of 41-60 years (11%) and a comparatively higher number of cases was recorded in age group 21-40 years (8.3%). Adults aged between 41 to 60 had a two times more likelihood to be infected with Tuberculosis than those aged below 20 years old. Moreover, pulmonary tuberculosis was highly prevalent among middle age (20-60) than any other age class in all study years (2014-2019). The pulmonary TB cases were highly predominant during the year 2014 which was 16.8% (19 of 113 subjects) whereas the almost a quarter of it (4.5%) was recorded in following year. Based on locality, the highest rate of infection was observed in Adobha (25%), a town at border of Sudan and Eritrea, in which patients who came from Adobha had 4 times more likelihood to be infected than those from Nakfa town.

**Conclusion:** This study showed Nakfa subzone has relatively increased prevalence of smear positive pulmonary tuberculosis than the average WHO estimate for the country. Therefore, appropriate policies and strategies for prevention, targeted detection of cases and treatment are required to reinforce Tuberculosis control programs.

## Introduction

Tuberculosis (TB) is one of the major infectious disease and health concern in the world. TB is a bacterial disease caused by Mycobacterium tuberculosis and infrequently by Mycobacterium africanum and Mycobacterium bovis [1]. TB pathogens are slow growing, fastidious, hydrophobic and lipid-rich bacteria that have acid fast rod shape which prevent decolorization with acid alcohol [2]. It is a chronic contagious infection that not only affects humans but a wide range of mammals. The main way of spread occurs by airborne transmission and infectious droplets [3]. A person with TB who is coughing is the key source of infection. The infectiousness of a person with TB disease is directly related to the number of tubercle bacilli that he or she expels into the air. Persons who expel many tubercle bacilli are more infectious than patients who expel few or no bacilli [3]. Since 2007, TB has been one of the top 10 causes of death, and the leading cause from a single infectious agent (Mycobacterium tuberculosis), ranking above HIV/ AIDS [2]. Globally, an estimated of 1.7 billion people are infected with M. tuberculosis and an estimated 10.0 million people (5–10%) fell ill with TB in 2018, a number that has been relatively stable in recent years [2]. Moreover, there were 1.2 million TB deaths among HIV-negative people in 2018 and an additional 251,000 deaths among HIV-positive people.

Geographically, most TB cases in 2018 were in regions of South-East Asia (44%), Africa (24%) and the Western Pacific (18%), with smaller shares in the Eastern Mediterranean (8%), the Americas (3%) and Europe (3%) [2]. Estimated tuberculosis incidence in the Horn of Africa ranges from 65 cases per 100,000 people per year in Eritrea to 192 cases per 100000 per year in Ethiopia, 274 cases per 100000 in Somalia, and 378 cases per 100,000 per year in Djibouti. Somalia has the highest estimated multidrug-resistant tuberculosis incidence, with 29 cases per 100,000 population per year [1].

Pulmonary Tuberculosis is primarily identified symptomatically using features like cough, fever, sweats, weight loss and hemoptysis and extra-pulmonary lymph nodeswelling (lymphadenitis) [3]. *A*lso disseminated/miliary TB is characterized by mycobacterial infection apart from lung and lymph node, in any part of the body, including the bone, meninges and kidneys [4]. The probability of developing TB disease is much higher among people living with HIV. It is also higher among people affected by undernutrition, diabetes, smoking and alcohol consumption [2].

The currently recommended treatment for cases of drug-susceptible TB disease is a 6-month regimen of four first-line drugs: isoniazid, rifampicin, ethambutol and pyrazinamide. The Global TB Drug Facility supplies a complete 6-month course for about US$ 40 per person [2]. Without treatment, the mortality rate from TB is high. In the absence of treatment with anti-TB drugs, a study found (conducted before drug treatments became available) that about 70% of individuals with sputum smear-positive pulmonary TB died within 10 years of being diagnosed, as did about 20% of people with culture-positive (but smear-negative) pulmonary TB [5]. Treatment for people with rifampicin-resistant TB (RR-TB) and multidrug-resistant TB (MDR-TB) is longer, and requires drugs that are more expensive (≥US$ 1000 per person) [2]. These medical intervention costs are significantly beyond the capacity of a country with a per capita GDP below US$ 200.

Multidrug resistant tuberculosis (MDR-TB) is defined as a disease caused by mycobacterium tuberculosis (TB) that is resistant to at least isoniazid and rifampicin, the two key first line drugs in short course TB chemotherapy [6]. Drug resistant tuberculosis is also a major threat in several countries and several measures have been integrated in their National Tuberculosis control programs. The latest data reported to WHO show a treatment success rate for MDR-TB of 56% globally [2].

The first-ever national anti-TB drug resistance survey was completed in Eritrea in 2018. According to this recent WHO estimate report, a total of 3100 new TB cases were present in 2018 in Eritrea which correspond to 89 cases per 100,000 population [7]. 140 of the cases were patients with TB and HIV coinfection and 66 of the cases had multidrug resistant TB. Nationally, 550 patients died from tuberculosis and related complications and 47 of them had HIV [7]. The 2018 survey also revealed that the incidence rate of estimated proportion of TB cases with MDR/RR-TB was 2% and 4.1% of them were from previously treated cases [7]. The problem of drug resistant TB exists in different parts of Eritrea but there is inadequate data on patterns of resistance among isolates of TB for specific drug type. Epidemiological research and analysis is in need of the time now for efficient preventive planning of TB control [8]. Very little information is available in Eritrea related to area specific prevalence of tuberculosis and risk factors attributed to it. This is a retrospective study aimed to assess the prevalence of tuberculosis disease among patients attending at Nakfa Hospital, Eritrea.

## Methods

### Study design and setting

The study was conducted at Nakfa Hospital in Nakfa subzone, 221km north of Asmara, capital of Eritrea between Latitude 16.6655 & Longitude 38.4768 from April 2014 to March 2019. Based on figures released by public health campaign programs in 2019, the Nakfa subzone comprises approximately 50,335 people living within its 10 administrative units. Most of inhabitants have a semi-nomadic way of life and are involved in grazing animals. This study was conducted to determine the prevalence of TB among the people on the basis of age, gender, address of patients and severity of pulmonary TB infection at Nakfa hospital. Sputum examination was carried out on every patient who was present with common symptoms of tuberculosis which is prolonged cough, chest pain, moderate fever, night sweat. Those who had positive sputum smear were considered as TB positive cases.

### Data Collection

The data for this research was extracted typically from secondary data source from already recorded documents including patient admission records and laboratory registers at Nakfa Hospital. Also all demographic details of subjects were retrieved from hospital records and were checked for completeness, cleaned manually and entered into SPSS statistical package.

### Statistical analysis

The data was analyzed statistically using IBM SPSS version 20. Descriptive statistics was used to evaluate the data and results were displayed in terms of tables, graphs & percentages. The chi-square test and fisher exact test was used to find the association between the variables like age and gender, and address of patients.

## Results

During the study period, a total of 1100 clinically suspected patients were subject to sputum exam at Nakfa hospital. The number of males and females was 475 (43.2%) and 625 (56.8%), respectively (Table 1). The result of the sputum examination by Acid fast staining showed that 86(7.8%), 95% CI: 6.4, 9.5%) subjects were present with smear positive pulmonary tuberculosis. The prevalence of Tuberculosis amongst males was 7.4% and 8.2% amongst females, which had no significant difference (*p* > 0.05). According to severity of infection, 38(3.5%) of subjects were +1 positive, whereas 23(2.1%) and 24(2.2%) of patients were +2 and +3 positives respectively. One of the subjects had actual number of acid fast bacilli (Figure 1). Moreover, most of subjects that visited the hospital were diagnosis cases (947[86.1%]) and rest were follow up cases (153[13.9%]).

**Table 1.** Demographic Characteristics of Sampled Subjects

| Variables |  | Percent distribution |  | Total |
| --- | --- | --- | --- | --- |
|  |  | Male N(%) | Female N (%) |  |
| Age group in years | < 20 | 123 (47.9) | 134 (52.1) | 257 |
|  | 20-40 | 125 (32.6) | 259 (67.4) | 384 |
|  | 41-60 | 157 (45.4) | 189 (54.6) | 346 |
|  | > 60 | 70(61.9) | 43 (38.1) | 113 |
| Exam Type | Diagnosis | 410(43.3) | 537(56.7) | 947 |
|  | Follow up | 65 (42.5) | 88(57.5) | 153 |
| Year | 2014 | 49 (43.4) | 64 (56.6) | 113 |
|  | 2015 | 79(39.5) | 121(60.5) | 200 |
|  | 2016 | 84(45.7) | 100(54.3) | 184 |
|  | 2017 | 99(43.6) | 128(56.4) | 227 |
|  | 2018 | 105(42.5) | 142(57.5%) | 247 |
|  | 2019 | 59(45.7) | 70(54.3) | 129 |
| Address | Nakfa | 242(42.8) | 324(57.2) | 566 |
|  | Meo | 39(42.4) | 53(57.6) | 92 |
|  | Beyan | 17(45.9) | 20(54.1) | 37 |
|  | Apollo | 39(60.9) | 25(39.1) | 64 |
|  | Afabet | 7(31.8) | 15(68.2) | 22 |
|  | Rora | 94(37.3) | 158(62.7) | 252 |
|  | Adobha | 16(66.7) | 8(33.3) | 24 |
|  | Agrae | 21(48.8) | 22(51.2) | 43 |

**Figure 1.**
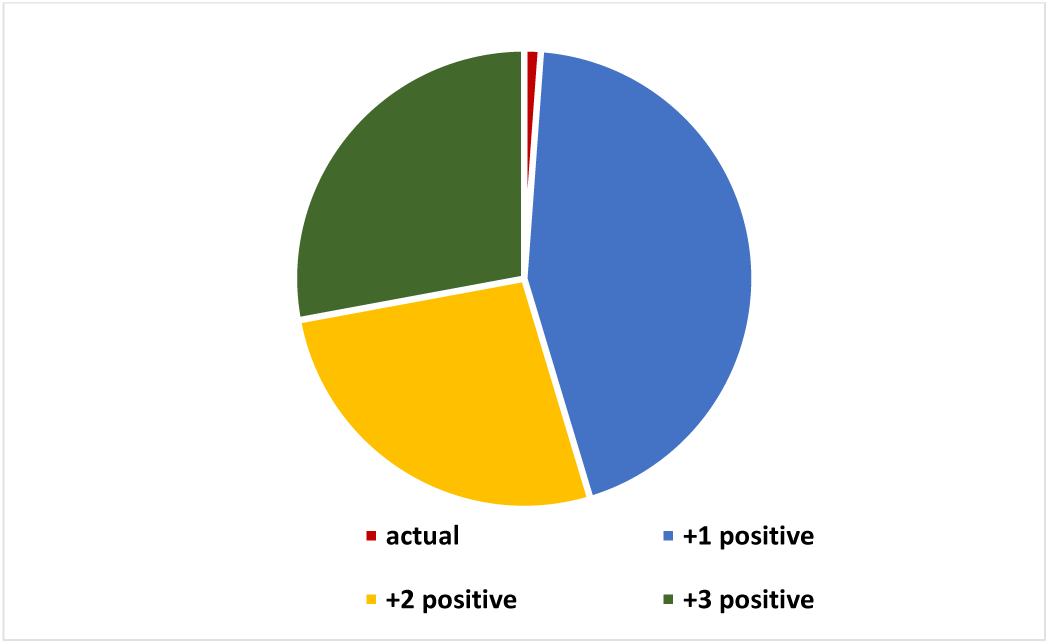
prevalence of pulmonary tuberculosis according to severity of infection, Nakfa hospital, Eritrea.

Age-wise analysis of the data shows that a statistically significant and comparatively higher number of cases (11.0%) was recorded in the age group 41 to 60 years and 8.3 % in age group 21 to 40 whereas the lowest percentages were observed in people aged from 1-20 years old (5.1%) and above 60 years old (2.7%). Binary logistic regression showed Adults aged between 41 to 60 had a more likelihood to be infected with Tuberculosis than those aged below 20 years old (OR: 2.32; 95%CI 1.21, 4.44) and subjects aged above 60 years old (OR: 4.52; 95%CI 1.36, 14.94). Moreover, pulmonary tuberculosis was highly prevalent among middle age (20-60) than any other age class in all study years (2014-2019) (Table 2).

**Table 2.** Prevalence of pulmonary tuberculosis amongst patients attending Nakfa Hospital, Eritrea (Apr. 2014 - March, 2019)

| Variables | Category | Total<br>N(%) | Pulmonary TB |  | COR [95%CI] | P- value |
| --- | --- | --- | --- | --- | --- | --- |
|  |  |  | Positive<br>N(%) | Negative<br>N(%) |  |  |
| Sex | Male | 475(43.2) | 35(7.4) | 440(92.6) | 1 | 0.63 |
|  | Female | 625(56.8) | 51(8.2) | 578(91.8) | 1.12(0.71, 1.75) |  |
| Age group in<br>years | < 20 | 257(23.4) | 13(5.1) | 244(94.9) | 1 | 0.01 |
|  | 20-40 | 384(34.9) | 32(8.3) | 352(91.7) | 1.70(0.88, 3.32) | 0.11 |
|  | 41-60 | 346(31.5) | 38(11) | 308(89) | 2.32(1.21, 4.44) | 0.01 |
|  | > 60 | 113(10.3) | 3(2.7) | 110(97.3) | 0.51(0.14, 1.83) | 0.30 |
| Year of infection | 2014 | 113(10.3) | 19(16.8) | 94(83.2) | 1 | 0.00 |
|  | 2015 | 200(18.2) | 9(4.5) | 191(95.5) | 0.23(0.10, 0.53) | 0.00 |
|  | 2016 | 184(16.7) | 21(11.4) | 163(88.6) | 0.63(0.32, 1.25) | 0.18 |
|  | 2017 | 227(20.6) | 11(4.8) | 216(95.2) | 0.25(0.12, 0.55) | 0.00 |
|  | 2018 | 247(22.5) | 13(5.3) | 234(94.7) | 0.28(0.13, 0.58) | 0.00 |
|  | 2019 | 129(11.7) | 13(10.1) | 116(89.9) | 0.55(0.26, 1.18) | 0.13 |
| Address | Beyan | 37 (3.4) | 1(2.7) | 36(97.3) | 1 | 0.02 |
|  | Apollo | 64 (5.8) | 5(7.8) | 59(92.2) | 3.05(0.34, 27.2) | 0.32 |
|  | Meo | 92 (8.4) | 3(3.3) | 89(96.7) | 1.21(0.12, 12.1) | 0.87 |
|  | Afabet | 22 (2.0) | 3(13.6) | 19(86.4) | 5.68(0.55, 58.4) | 0.14 |
|  | Rora | 252 (22.9) | 24(9.5) | 228(90.5) | 3.79(0.49, 28.8) | 0.19 |
|  | Adobha | 24 (2.2) | 6(25) | 18(75) | 12(1.34, 107.3) | 0.03 |
|  | Agrae | 43 (3.9) | 6(14) | 37(86) | 5.83(0.67, 50.9) | 0.11 |
|  | Nakfa | 566 (51.5) | 38(6.7) | 528(93.3) | 2.59(0.35, 19.4) | 0.35 |
| Seasons | Summer | 252(22.9) | 16(6.3) | 236(93.7) | 1 | 0.02 |
|  | Autumn | 205(18.6) | 27(13.2) | 178(86.8) | 2.24(1.17, 4.28) | 0.01 |
|  | Winter | 332(30.2) | 22(6.6) | 310(93.4) | 1.04(0.53, 2.03) | 0.88 |
|  | Spring | 311(28.3) | 21(6.8) | 290(93.2) | 1.07(0.54, 2.09) | 0.84 |

A statistically significant severe tuberculosis (3+ positive) was present more persistently in females (3.2% vs 0.8%) whereas common 1+ and 2+ positive AFB smear results were seen in males with 3.8% vs 3.2% and 2.7% vs 1.6% percent respectively. Similarly, a significant difference was observed between year of infection and positive AFB smears (p-value=0.00). Figure 2 demonstrates the pattern of prevalence of pulmonary TB across all six study years. The pulmonary TB cases were highly predominant during the year 2014 which was 16.8% (19 of 113 subjects) whereas the almost a quarter of it (4.5%) was recorded in following year. This difference was found to be statistically significant. Also a comparable prevalence of pulmonary TB disease was seen during 2016 (11.4%) and in 2019 (10.1%). Moreover, the prevalence of pulmonary TB was observed in statistically significant difference among different seasons (p-value=0.018). Compared with others, Autumn had the highest prevalence rate (13.2%) with rest seasons having comparable percentages.

**Figure 2.**
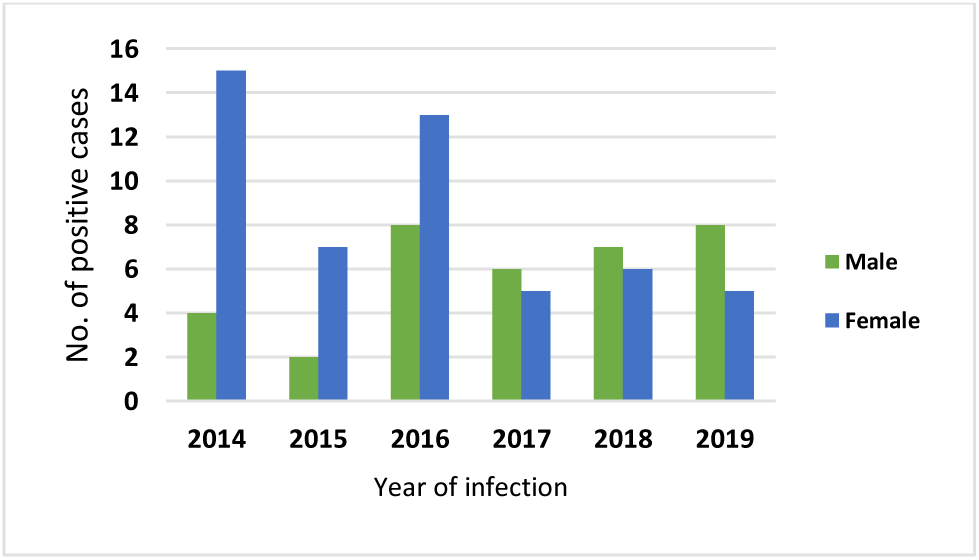
The prevalence of pulmonary TB disease among males and females from 2014-2019 at Nakfa Hospital, Eritrea.

More than half (51.5%) of the subjects who had visited Nakfa Hospital resided inside Nakfa town whereas the rest were from small towns far around. The highest rate of infection was observed in Adobha (25%), a small town at the border of Sudan and northern Eritrea. In binary logistic regression, patients who came from Adobha had 4 times more likelihood to be infected than those from Nakfa area (OR: 4.63; 95%CI 1.73, 12.4). Also comparable prevalence was observed in Agrae (14%) and Afabet (13.6%) areas. The lowest prevalence was generally observed in Nakfa town and its periphery areas. The difference was statistically significant (p-value= 0.008) (Table 2).

## Discussion

TB is the one of the major causes of death in persons aged 15-49 years and the primary cause from a single infectious agent (Mycobacterium tuberculosis) with an estimated 10 million cases and 1.2 million deaths as of 2018 [2]. It is also estimated that almost one third of the world population harbor the causative agent of TB in their lungs. Demographic characteristics analysis of this study reveal female subjects had a higher distribution among the sexes [625(56.8%) out of 1100]. Age-wise analysis also shows age group 20 to 40 had the highest portion of study participants [384(35%) out of 1100]. The result of current study shows pulmonary TB prevalence of 7.8% (86 out of 1100) subjects within the study time frame. As compared to previous survey done in Eritrea, this finding is higher than WHO report and range for prevalence reported in Eritrea by world organizations [7]. However, the prevalence of TB was lower compared to studies conducted in different parts of horn of Africa [9, 10, 11].

The prevalence of pulmonary TB was higher amongst females (8.2%) compared to males (7.4%) which had no significant difference (*p* > 0.05). But it is reported that females are more prone to development of disease as they are more immune deficient [12]. This finding is in agreement with previous studies conducted in different parts of the world [13, 14].

The prevalence of sputum smear-positive TB in this study showed a statistically significant and comparatively higher number of cases (11.0%) in the age group 41 to 60 years and 8.3% in age group 21 to 40. However, the lowest percentages were observed in people aged from 1-20 years old (5.1%) and above 60 years old (2.7%). These findings are in line with the results of others researches which indicated a large number of cases prevalent in the creative and economically important age groups [15]. Different studies also indicated differences in prevalence of smear positive TB among various age categories [9, 16, 17]. The result is also consistent with WHO report indicating the highest burden of TB in adult men accounting for 57% of all TB cases in 2018 [2].

Similarly, a significant difference was observed between year of infection and positive AFB smears (p-value=0.00). The pulmonary TB cases were highly predominant during the year 2014 which was 16.8% (19 of 113 subjects) whereas the almost a quarter of it (4.5%) was recorded in following year. This steep decline of TB prevalence was attributed to continuous control efforts implemented in the study area. However, further investigation should be done as the rate increased to 10% in recent couple of years. Prevalence of pulmonary TB was observed in statistically significant difference among different seasons (p-value=0.018). Compared with others, Autumn had the highest prevalence rate (13.2%) with rest seasons having comparable percentages (Table 2).

This differences may be related to variations in food availability and intake, Vitamin D level variability, indoor activities, seasonal change in immune function, and delays in the diagnosis and treatment of tuberculosis being potential causes of seasonal tuberculosis disease [18].

## Conclusion

Eritrea is one of the few countries that have achieved Millennium Development Goals (MDGs) in the health sector in general and in communicable disease transmission in particular.

This study showed Nakfa area has relatively increased prevalence of smear positive pulmonary tuberculosis than the average WHO estimate for the country. Therefore, appropriate policies and strategies for prevention, targeted detection of cases and treatment are required to reinforce TB control programs. It should be noted that, compared to others, the low prevalence finding in this study may be attributed to methodology difference. In this study, subjects were screened using Ziehl-Neelsen acid fast staining technique in direct smear which has lower sensitivity compared to bleach concentrated smears.

## Data Availability

The data used to support the findings of this study are available from the corresponding author upon request.

## Abbreviations

TB: Tuberculosis
M.tb: Mycobacterium tuberculosis
WHO: World health Organization
MDR-TB: Multidrug resistant Tuberculosis
RR-TB: Rifampsin resistant Tuberculosis

## Ethical approval and consent to participate

The data was collected after ethical clearance was obtained from the Department of Clinical Laboratory Science, Asmara College of Health Sciences. After discussing the objective and methods of the study, a written consent was acquired from Nakfa Hospital before the data collection period. In order to maintain anonymity and ethical confidentiality, all patients were given codes during the data extraction process and patient names were not extracted from the register.

## Consent for publication

Not applicable.

## Funding

No source of funding was needed to carry out this study.

## Competing interests

The author declare that no competing interests exist.

## Acknowledgment

The author extremely would like to acknowledge Nakfa Hospital officials and study participants throughout the study years.

## Notes

### Competing Interest Statement

The authors have declared no competing interest.

